# Can AI Match Human Experts? Evaluating LLM-Generated Feedback on Resident Scholarly Projects

**DOI:** 10.64898/2026.03.04.26346878

**Authors:** Zack van Allen, Sylvie Forgues-Martel, Maddie J. Venables, Yosr Ghanney, Alexandre Villeneuve, Jarvis Dongmo, Meherin Ahmed, Douglas Archibald, Kheira Jolin-Dahel

**Author notes:** Corresponding author: Dr. Kheira Jolin-Dahel.

## Abstract

**Background:** Delivering timely, high-quality feedback on resident scholarly projects is labour-intensive, especially in large programmes. We developed an AI-assisted evaluation system, powered by the open-weight LLaMA-3.1 large-language model (LLM), to generate formative feedback on Family Medicine residents’ scholarly projects and compared its performance with expert human evaluators.

**Methods:** We evaluated whether the AI-generated feedback achieves comparable quality to expert feedback. The tool ingests heterogeneous resident submissions (PDFs, scans, photographs) via OCR and produces section-by-section feedback aligned with programme rubrics. In a three-phase study we evaluated 240 feedback reports (Short, Question and Timeline, Final; n = 80 each). Within each phase, 40 reports were AI-generated and 40 produced by research experts across four project types: Quality Improvement, Survey-Based, Research, and Literature Review. Blinded raters used a 25-item survey across five constructs: understanding & reasoning, trust & confidence, quality of information, expression style & persona, safety & harm.

**Results:** Survey reliability was high across phases (α = .71–.98). Human feedback generally out-scored AI. In short reports, humans led on quality (Mean ± SD; 4.14 ± 0.57 vs 3.09 ± 1.05) and trust (3.96 ± 0.71 vs 2.78 ± 1.15). In final reports, differences become small for quality (4.09 ± 0.65 vs 3.49 ± 0.68) and persona (4.16 ± 0.40 vs 3.91 ± 0.50), while AI was preferred for safety (4.50 ± 0.60 vs 4.36 ± 0.56). Performance varied by project type: in survey-based final reports the AI led on quality (4.28 ± 0.50 vs 3.98 ± 0.44) and safety (4.58 ± 0.40 vs 4.24 ± 0.67), whereas in quality-improvement short reports humans were markedly superior in reasoning (4.27 ± 0.68 vs 2.33 ± 1.00).

**Conclusions:** An open-weight LLM with curated prompts can generate rubric-aligned feedback at scale that approaches the quality of expert human feedback. While expert feedback remained superior overall, AI surpassed humans in selected contexts and safety assessments. Performance of the tool will increase over time as newer and more capable open-weight models are released. Our code and systems prompts are open source.

---

All postgraduate programmes are required to learn about and participate in research (College of Family Physicians of Canada [CFPC], 2018) and timely, high-quality feedback is critical for developing research proficiency (Shafian et al., 2024; Burgess, van Diggele, Roberts, & Mellis, 2020), yet large residency programmes struggle to provide this at scale. At the University of Ottawa, the Family Medicine Resident Scholarly Project (FMRSP) is a two-year scholarly project requirement in our Family Medicine residency, intended to cultivate research skills and critical thinking. Approximately 170–180 residents are at various project stages in a given year, each expecting detailed feedback at three key milestones (proposal, short report, final report). This volume of reviews has stretched our support staff’s capacity, and feedback turnaround can exceed 60 days for some stages. Such delays hinder residents’ progress and add pressure as deadlines approach.

An expanding body of work in medical education suggests that AI-generated feedback can be effective across various training levels, with generally positive reception and performance approaching human evaluators in structured tasks. For example, ChatGPT feedback was rated as timely and helpful by second-year medical students in Korea (Park, 2023), and GPT-3.5 even outscored standardized-patient raters on clinical note assessments in a U.S. study (Burke et al., 2024). Similarly, GPT-4 variants have shown promise: GPT-4V provided relevant item-level feedback on progress tests (Tomova et al., 2024), and GPT-4’s scoring of clinical reasoning notes correlated strongly with expert ratings (Schaye et al., 2025). AI has also been integrated into teaching interventions, interns who used ChatGPT during problem-based learning improved their knowledge and evaluation scores (Zeng et al., 2025) and the model can generate assessment items (vignettes and MCQs) with human-comparable quality (Coşkun et al., 2024)

Controlled trials provide a more nuanced picture. In a single-centre RCT (n = 129), GPT 3.5 feedback on clinical reasoning questions yielded learning gains similar to expert comments overall, though experts remained superior for complex urinary-tract scenarios (Çiçek et al., 2025). Conversely, a double-blind RCT using AI patient simulations found that four sessions of GPT 3.5 generated formative feedback significantly improved clinical-decision-making scores compared with simulation alone (Brügge et al., 2024). However, not all findings are uniformly positive – some RCTs show AI feedback matching expert performance or improving learner outcomes, while others show no benefit in complex tasks (Çiçek et al., 2025; Brügge et al., 2024; Goh et al., 2024). This inconsistency underscores the need for further evaluation of AI feedback tools in authentic residency education settings. Despite rapid growth of this literature, few studies have evaluated AI feedback within postgraduate medical education or used open-weight models. Finally, early procedural work, such as editorial evidence on automated feedback during central-venous-catheterization training, highlights parallel advances in skills instruction (Tsai et al., 2025). It is noteworthy that, due to the rapid pace of AI development and the slower cycle of academic publication, many of these studies rely on now-deprecated models (e.g. GPT-4 and earlier). Nonetheless, collectively these studies suggest that AI-driven feedback can approximate human evaluators on certain cognitive tasks and significantly accelerate formative feedback cycles.

To address the challenge of providing quality feedback at scale, we developed an AI-assisted feedback tool for resident scholarly and research projects, using an open-weight large language model deployable at any institution. Our objective was to evaluate whether such a model could generate rubric-aligned, high-quality and safe feedback comparable to that of human experts while substantially reducing review time. This tool is designed to provide research feedback of sufficient quality to be indistinguishable from feedback generated by a (human) research expert. The tool uses an open-weight LLM that can be locally deployed in any program; its model-agnostic design means medical educational professionals can easily swap in more capable models as they emerge, future-proofing the system’s performance.

Here, we describe the development of our open-weight AI-driven feedback system and an initial evaluation of its performance against human generated feedback in our Family Medicine residency, examining whether it can deliver high-quality, safe feedback more efficiently at scale.

## Methods

This study evaluated feedback reports generated by both AI and expert human evaluators across three distinct project phases: short reports (SR), question and timeline reports (QT), and final reports (FR). A total of 240 feedback reports were assessed, with 80 reports for each phase (SR, QT, FR). In each phase, half of the reports (n=40) were generated by AI and half (n=40) by an expert human evaluator. The study took place between Aug 2024 and Feb 2025. The feedback was generated for four types of projects: quality improvement (QI), survey-based (SB), research-based (RB), and literature review (LR), with an equal number of reports per type in each phase. Four experienced researchers evaluated these reports using a 25-item survey adapted from Tam et al (2024) assessing five constructs: understanding and reasoning, trust and confidence, quality of information, expression style and persona, and safety and harm. The question stem asked participants to ‘please state your level of agreement or disagreement with the following statements based on the quality of the generated feedback’ on a scale from 1 (strongly disagree) to 5 (strongly agree). This project falls under TCPS2 Article 2.5 and, as such, did not require Research Ethics Board (REB) review, as confirmed by the University of Ottawa REB during an initial review for exemption and reported on August 29, 2024.

To prepare the AI tool, our methodological approach was divided into three main stages: data preprocessing and extraction, model selection and prompt design, and interface development for evaluator interaction.

### Data Preprocessing and Extraction

The initial phase involved collecting and preprocessing resident reports submitted in highly heterogeneous formats, including digitally typed PDFs, handwritten scans, and photographs of printed forms. This variability posed significant challenges for reliable content extraction. Multiple extraction techniques were tested, including native PDF parsers such as PyMuPDF (PyMuPDF Development Team, 2025), PDFPlumber (Singer-Vine, 2025), and pdfrw (Maupin, 2021). These libraries extracted PDF form inputs with their associated labels and any additional text. To enhance performance, we also tested table-recognition libraries using OpenCV (OpenCV Development Team, 2024) and document-layout analysis tools. However, because of inconsistent document structure and quality, optical character recognition (OCR) proved most robust. Tesseract OCR (Tesseract OCR Development Team, 2025), enhanced with layout-aware parsing, delivered superior performance in accurately extracting textual content, figures, and tables compared with traditional PDF-parsing methods.

Despite these advances, figures and tabular data remained particularly difficult to extract consistently. To address this, we explored the use of Large Vision Models (LVMs) for semantic image interpretation. However, empirical evaluation revealed a high rate of hallucinations and inconsistency in figure-text alignment. Consequently, the extraction pipeline remained centered on OCR-based processing. Extracted content was programmatically structured and exported into Excel spreadsheets, with schema-specific formatting for each report category. Each Excel file reflected a standardized mapping of document sections (e.g., Introduction, Methodology, Results, Discussion), ensuring consistent downstream input for the AI evaluation engine. The quality of extracted text was evaluated to verify accuracy.

### AI Model Selection and Prompt Design

Following data preprocessing, the second phase focused on designing the AI model responsible for evaluating the reports and generating formative feedback. In selecting a suitable large language model (LLM), key considerations included performance, data privacy, and the ability to operate in a controlled local environment. We selected LLaMA 3.1, an open-source model that could be deployed securely and customized for our institutional needs. Future deployments could select any open-weight (or commercial) model.

Our initial implementation employed a zero-shot prompting approach, wherein the model was instructed to evaluate reports based solely on descriptive prompts aligned with pedagogical rubrics. However, this approach yielded suboptimal coherence and lacked alignment with human evaluations. To address this, we adopted a *few-shot learning* paradigm. This involved curating exemplar pairs of student submissions and evaluator feedback to condition the model’s responses through in-context learning. Incorporating these exemplars into the prompt significantly improved the model’s ability to emulate evaluation styles and provide pedagogically relevant feedback. Each prompt was tailored to the report category and included both structural guidance (e.g., required sections, evaluation criteria) and stylistic targets (e.g., formative tone, specific domain vocabulary).

### Evaluation Interface and Feedback Generation

To facilitate real-world use by faculty evaluators, we developed a web-based application that integrates the AI evaluation pipeline. The interface allows evaluators to select the report type and category, upload the PDF file, and initiate an AI-based evaluation. The output is generated in a Word document format, containing section-by-section feedback aligned with educational rubrics. This output is editable, enabling evaluators to revise or augment the AI-generated content before finalizing their feedback. This human-in-the-loop design ensures both efficiency and academic oversight, preserving the evaluator’s role in assessment while leveraging the AI’s ability to rapidly process and interpret structured input. This web-based application was deployed with Amazon AWS. We created a repository for the Web application frontend with HTML, CSS and JavaScript which was then deployed using AWS Amplify. We also used Lambda functions and Batch from AWS to preprocess the data, use the AI model with a performant GPU instance and then post process the generated text into a Word document.

## Results

### Measurement Reliability

We examined one form of reliability, internal consistency of the survey items within each report type, using Cronbach’s α. Cronbach’s α indicated good-to-excellent internal consistency across all report stages (Table 1). Reliability was uniformly high for short reports and question and timeline reports (α ≥ .83), while final reports remained acceptable overall, though lowest for expression style & persona (α = .71). Dropping a single item improved safety & harm reliability in question and timeline reports from .70 to .85, confirming the survey’s robustness after minor refinement.

**Table 1:** Summary of Cronbach’s Alpha Scores for FR, QT, and SR.

| Construct | Question and Timeline<br>(QT) | Short Reports<br>(SR) | Final Reports<br>(FR) |
| --- | --- | --- | --- |
| Quality of Information | .96 | .98 | .96 |
| Understanding & Reasoning | .93 | .96 | .85 |
| Expression Style & Persona | .83 | .96 | .71 |
| Safety & Harm | .85† | .92 | .91 |
| Trust & Confidence | .91 | .92 | .86 |

† For the QT "Safety & Harm" construct, dropping one survey item improved Cronbach's $\alpha$ from .70 to .85.

### Overall Comparison of Human and AI-Generated Feedback

Table 2 shows a clear favour of expert reviewers, but the magnitude of advantage shrinks for final reports. Descriptive results are presented as Mean ± SD. Across all five quality dimensions, human scores clustered tightly between 4.09 ± 0.65 and 4.75 ± 0.51, whereas AI scores spanned a broader 2.78 ± 1.15 to 4.50 ± 0.60 range. The largest discrepancies appeared in short reports: human reviewers out-performed the model by 1.05 points on quality (4.14 ± 0.57 vs 3.09 ± 1.05), 0.97 on reasoning (4.23 ± 0.68 vs 3.26 ± 1.22), and 1.18 on trust (3.96 ± 0.71 vs 2.78 ± 1.15). For question and timeline reports, gaps narrowed but remained pronounced for trust (4.21 ± 0.64 vs 3.28 ± 0.78) and quality (4.36 ± 0.51 vs 3.69 ± 0.70). In final reports the two systems converged: reasoning was effectively equal (4.07 ± 0.53 vs 4.01 ± 0.55), and AI slightly surpassed humans on safety (4.50 ± 0.60 vs 4.36 ± 0.56), indicating the model’s most consistent strength. Overall, human feedback was more precise (smaller SDs) and dependable, whereas AI feedback exhibited greater variability, particularly in early-stage work.

**Table 2.** Average ratings for AI and Human feedback.

| Dimension | Question and Timeline |  |  | Short Report |  |  | Final Report |  |  |
| --- | --- | --- | --- | --- | --- | --- | --- | --- | --- |
|  | AI<br>M (SD) | Human<br>M (SD) | Cohen's<br>d | AI<br>M (SD) | Human<br>M (SD) | Cohen's<br>d | AI<br>M (SD) | Human<br>M (SD) | Cohen's<br>d |
| Quality | 3.69<br>(0.70) | 4.36<br>(0.51) | 1.10 | 3.09<br>(1.05) | 4.14<br>(0.57) | 1.26 | 3.49<br>(0.68) | 4.09<br>(0.65) | 0.21 |
| Reasoning | 3.56<br>(0.88) | 4.23<br>(0.66) | 0.86 | 3.26<br>(1.22) | 4.23<br>(0.68) | 0.99 | 4.01<br>(0.55) | 4.07<br>(0.53) | 0.11 |
| Persona | 3.54<br>(0.63) | 4.21<br>(0.61) | 1.07 | 3.52<br>(1.08) | 4.20<br>(0.62) | 0.77 | 3.91<br>(0.50) | 4.16<br>(0.40) | 0.55 |
| Safety | 4.27<br>(0.74) | 4.75<br>(0.51) | 0.75 | 3.79<br>(0.95) | 4.28<br>(0.70) | 0.59 | 4.50<br>(0.60) | 4.36<br>(0.56) | -0.24 |
| Trust | 3.28<br>(0.78) | 4.21<br>(0.64) | 1.32 | 2.78<br>(1.15) | 3.96<br>(0.71) | 1.24 | 3.73<br>(0.72) | 3.80<br>(0.85) | 0.10 |

Note: M = mean, SD = standard deviation, Cohen's d conventions: small ( $d = .2$ ), medium ( $d = .5$ ), large ( $d = .8$ ). Quality = Quality of Information, Reasoning = Understanding & Reasoning, Persona = Expression Style & Persona, Safety = Safety & Harm, Trust = Trust & Confidence.

### Feedback Ratings by Case Type

Performance patterns shifted when results were broken down by project genre (Table 3, Figure 1). In survey-based final reports, AI edged out experts on four of five dimensions; its quality score reached 4.28 ± 0.50 versus 3.98 ± 0.44 for humans, and its safety rating peaked at 4.58 ± 0.40 compared with 4.24 ± 0.67, suggesting the model handles structured, data-centric projects well. Conversely, experts maintained a modest lead in narrative-heavy literature review final reports, posting higher means for quality (3.85 ± 0.87 vs 3.45 ± 0.61) and persona (4.17 ± 0.39 vs 3.70 ± 0.29), though AI again delivered safer comments (4.42 ± 0.89 vs 4.02 ± 0.47). The starkest contrast emerged in quality-improvement short reports, where sparse, context-specific material magnified AI’s weaknesses: reasoning plunged to 2.33 ± 1.00 and trust to 2.25 ± 1.06, while human reviewers remained solidly above four on both metrics (4.27 ± 0.68 and 3.95 ± 0.64, respectively). Across genres, AI’s most reliable dimension was safety, whereas trust and quality consistently lagged, especially in complex, early-stage work, highlighting the continued pedagogical value of expert oversight in those contexts.

**Figure 1.**
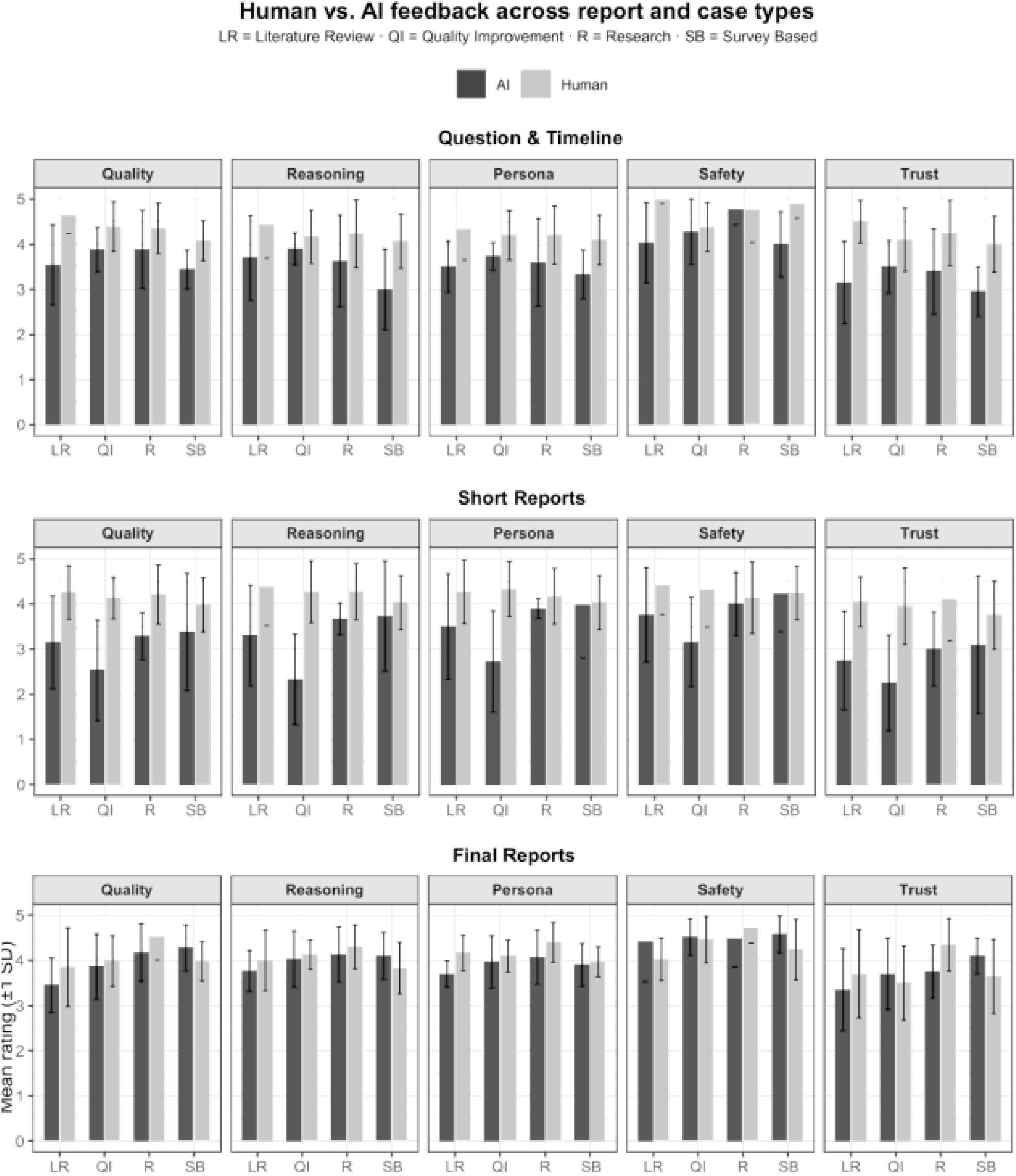
Human vs AI feedback across report and case types

**Table 3.**
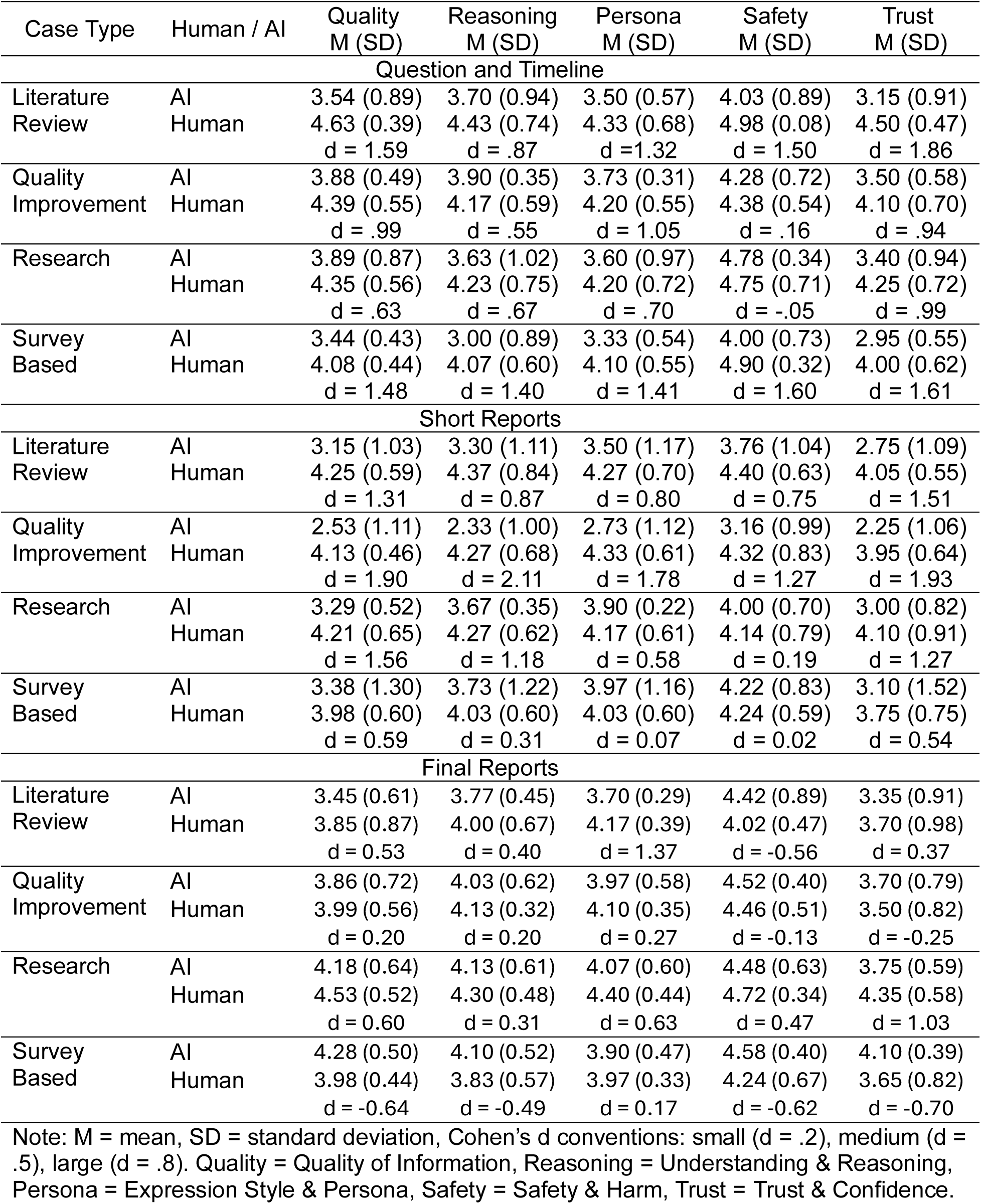
Ratings by case type and human vs AI feedback.

## Discussion

We evaluated a custom open-weight AI tool designed to generate formative feedback on Family Medicine resident scholarly and research projects. In a head-to-head comparison of 240 feedback reports (Short, question and timeline and Final report stages) spanning four project genres, human reviewers still outperformed the AI on most dimensions, especially *trust/confidence* and *expression style*. However, the performance gap decreased over time (from short reports to final reports) reflecting improvements in the AI system (e.g., the refinement of prompts and data extraction methods over time). Differences in final reports were generally small to medium, and the AI even marginally outscored human feedback on safety/harm in final reports. These results suggest that structured prompt engineering and iterative training can progressively align AI-generated feedback with human expert standards, and echo broader evidence that LLM feedback can approach human quality on structured cognitive tasks while accelerating review cycles (Burke et al., 2024; Tomova et al., 2024; Schaye et al., 2025).

Timed, specific critique is one of the strongest pedagogical levers in research training (Hattie & Timperley, 2007; Burgess et al., 2020; Shafian et al., 2024), yet our program’s traditional turnaround exceeded 60 days for some milestones due to resource constraints coupled with a high volume of research projects. We did not formally measure expert rater time burden, so whether AI-assisted reviews reduce effort compared with traditional feedback remains unknown and warrants prospective evaluation. However, our AI tool reduced the delay of processing a feedback request to several minutes for feedback generation, enabling rapid iteration while a human-in-the-loop approach (Mosqueira-Rey et al., 2023) was used to endorse or refine comments. This hybrid model can also promote equity and consistency in feedback – every resident receives a core level of rubric-based critique regardless of variable human input – while allowing supervisors to focus their efforts on nuanced mentorship rather than repetitive comments or copy-editing.

Another key consideration is the accuracy and fairness of the AI’s comments. LLMs can occasionally hallucinate misinformation or reflect biases present in their training data. Reassuringly, our evaluation found no egregious safety issues – in fact, the AI’s feedback was rated slightly *safer* (i.e. more free of harmful or inappropriate content) than human feedback in certain instances. This likely reflects the model’s neutral, polite, non-judgmental style and adherence to rubric criteria. However, the absence of red flags in our sample does not eliminate the risk. Ongoing human oversight remains essential to catch subtle errors or biases that the model might introduce. Until data shows AI feedback is errorless, we recommend that a faculty member or senior reviewer always vet the AI-generated feedback before it is released to residents, maintaining accountability and serving as quality control. This not only prevents the occasional hallucination from reaching the learner but also reinforces the accountability of the feedback process.

Careful consideration should also be given to the pedagogical trade-offs of providing AI enabled feedback to residents compared to teaching learners how to reliably obtain feedback on their own using AI tools (Abdulnour 2025; Bearman & Ajjawi 2023; Boscardin 2024). As researchers and educators continue to refine prompting strategies and test model performance, we should aim to validate the combination of prompting strategies and AI models that results in sound academic feedback and share those techniques directly with learners. Also, future work should empirically test how AI feedback influences residents’ metacognition and self-regulated learning behaviours. Beyond efficiency gains, however, there is also a risk that over-reliance on AI may encourage surface-level engagement or even reduce the effort learners put into self-directed improvement (Yu 2024; Jabbour 2023). To address this, training should not only expose residents to AI-generated comments but also teach them to critically appraise and cross-check those comments against rubrics and established scholarly standards using evidence-based steps and a “verify-then-trust” habit (Abdulnour 2025). Embedding this form of AI literacy into residency education ensures that learners gain both the immediate benefits of timely, structured feedback and the long-term capacity to use AI responsibly, as one tool among many in their academic development. In other words, the next frontier is not letting AI teach for us, but teaching clinician scholars to think with AI. The real wins are not limited to faster feedback alone, but include more equitable access to timely critique, stronger development of residents’ research skills, and cultivating graduates who can use AI wisely and confidently as part of their scholarly toolkit.

### Limitations and Future Directions

Initial results demonstrate that a customized, open-source AI assistant can provide useful, rubric-aligned feedback at scale, narrowing – though not yet closing – the gap with expert reviewers. However, our approach has limitations. The current model proved underwhelming in early-stage Short Reports, where sparse content led to generic comments that inspired less trust than expert feedback.

Additionally, the AI model struggled with Quality Improvement (QI) projects, which often involve local system processes and context-specific interventions. The model sometimes failed to appreciate the practical nuances of QI proposals, resulting in feedback that missed the mark. This pattern of *context-dependent* performance aligns with findings elsewhere in the literature with studies finding AI-generated feedback to be on par with experts in some settings but notably worse in others, especially for complex, domain-specific problems (Çiçek et al., 2025; Goh et al., 2024). For instance, ChatGPT’s teaching feedback has been shown to be comparable to an expert’s on average, yet inferior when the case involved a complicated scenario requiring specialized knowledge (Çiçek et al., 2025). Likewise, Goh et al. (2024) reported no significant benefit from LLM assistance in a nuanced diagnostic reasoning task. Our results mirror these trends, suggesting that while the evaluated AI models may handle generic research tasks well, they are less reliable for projects that require rich contextual awareness. Beyond content-related limitations, there are methodological caveats: because this study was conducted in a single institution and within one family medicine residency program using a locally developed review process, the generalizability of our findings to other programs or specialties is unknown and will require replication in other settings. Although we attempted to blind evaluators, it’s possible they picked up on stylistic differences between AI and human feedback, introducing bias. Moreover, we assessed feedback quality and safety, but not downstream learning outcomes – whether receiving AI-generated comments actually helps residents improve their projects or skills remains an open question for future research. Future studies may benefit from explicitly incorporating resident outcomes (e.g., improvement in subsequent drafts, rubric score progression or perceptions of feedback utility) to determine the educational impact of AI-assisted reviews.

Importantly, the system uses a model-agnostic architecture (LLaMA 3.1 in the present study) that can be exchanged for stronger open-weight models as they appear, preserving data privacy while future-proofing performance. Since deploying the tool across an academic year, the performance of open-source models have increased. We expect newer models to demonstrate enhanced ability to provide research project feedback, especially models with reasoning capabilities. We recommend using the strongest open-weight model.

## Conclusion

In conclusion, our initial results demonstrate that a tailored AI feedback assistant can deliver useful, rubric-aligned critiques at scale, partially bridging, though not yet closing, the gap between machine and expert feedback. There are multiple benefits of AI feedback, beyond timeliness, which has been a longstanding pain point in our residency program. It can promote more equitable access to critique, support deeper scholarly skill-building and can help residents develop the judgment to use AI responsibly. At the same time, it underscored the enduring value of human judgment for complex, context-rich tasks and the importance of a synergistic human–AI approach. We are encouraged that the platform’s performance will only grow as more capable models emerge and as we learn how best to integrate them. Future studies should examine whether AI feedback translates into measurable improvements in resident project quality and skill development, time saving for faculty and how best to embed AI literacy training into residency curricula. With continued refinement, rigorous evaluation, and mindful implementation, such human–AI partnerships could enable medical educators to provide timely, equitable feedback to all learners at scale without compromising quality or ethics. A final consideration is the need for periodic recalibration of the system as open-weight models evolve, ensuring reproducibility, stability and alignment with shifting assessment standards.

## Data Availability

Code will be made available following publication

## Appendix I Item-Level Analysis

Item-level difference scores (Human - AI) revealed context-specific patterns of comparative strength across the four case types and five dimensions. Positive values indicate human superiority and negative values indicate AI superiority.

For **Final Reports (FRs; Table 4)**: Literature review items generally showed modest positive scores (human advantages), particularly for Quality and Understanding (e.g., ESL6 Understanding =1.00; ESL4 Quality=1.83). Safety often favored AI (e.g., ESL3 =−1.60). Trust differences were mostly slight, except for a large human edge on ESL4 (3.00). Quality improvement items showed heterogeneous differences. Humans led on about half, especially ESQ7 and ESQ10 (Quality 0.92; Persona 1.00-1.33). AI surpassed humans on multiple Safety and Trust scores (e.g., ESQ3 Safety =−0.40, Trust =−1.50). Research report items displayed the widest spread. Dramatic human superiority occurred in ESR3 (Quality =2.00; Safety =1.80; Trust =2.00). AI outperformed on ESR8 across four dimensions (e.g., Quality =−1.08; Understanding =−1.33). Survey items showed consistent, though moderate, AI advantages for Quality and Understanding (e.g., ESS2 Quality =−1.00). Human strengths emerged sporadically (e.g., ESS6 Trust =1.00; Persona =1.00). Safety had the largest absolute differences, from a strong human edge (ESS5 0.60) to the greatest AI lead (ESS9 =−1.60). These swings underscore the underlying variability in the model performance that aligns with genre complexity and structure. Across dimensions in FRs, Safety generated the most extreme swings (Human lead 1.80 to AI lead 1.60). Trust differences were typically smaller but could be pronounced (ESL4 3.00).

For **Question and Timeline Reports (QTs; Table 4)**: Literature review items (EQL) predominantly showed positive difference scores (human superiority), though some instances favored AI (EQL3, EQL7 in Quality and Understanding). EQL9 showed substantial human advantages (Quality: 2.67, Trust: 2.50). Quality improvement items (EQQ) were heterogeneous. Many showed moderate human advantages, but AI performed better in Safety for several cases. EQQ8 showed consistent AI advantages, especially in Trust (−1.50). Research report items (EQR) had the most variable pattern. EQR2 showed pronounced human advantages (Understanding: 4.00, Trust: 3.50), while EQR4 and EQR9 showed consistent AI advantages. Survey items (EQS) most consistently favored human feedback. Safety ratings had particularly strong human advantages (several ≥1.50). To summarize, humans were rated higher in Trust and Quality, while AI occasionally excelled in Safety, particularly for structured project types. Across dimensions in QTs, Safety showed the highest variability (Human lead 2.25 to AI lead −1.75). Trust generally favored humans, with exceptions.

For **Short Reports (SRs; Table 6)**: Literature review items (ESL) had considerable variability (range: −0.67 to 3.08). ESL7 (Quality: 3.08) and ESL2 (Understanding: 3.67) showed high human-AI disparities. ESL5 suggested superior AI performance (differentials −1.60 to 0.33). Quality improvement items (ESQ) predominantly showed positive differentials (human superiority). ESQ3 had consistently high positive differentials (Quality: 2.75, Trust: 3.00), while ESQ9 showed neutral differentials. Research report items (ESR) showed more moderate differentials. ESR7 (Quality: 2.17) and ESR10 (Safety: −2.60) were notable. Safety had high variability here (−2.60 to 1.60). Survey items (ESS) had the most pronounced range of differentials (−2.00 to 3.00). ESS4 showed high positive differentials (Understanding: 3.00, Persona: 3.00), while ESS8 showed uniformly negative differentials (AI superiority, range −2.00 to −1.83). Across dimensions in SRs, Quality metrics consistently showed positive differentials (human superiority). Trust differentials were more stable. Safety evaluations exhibited the highest variability, especially in Research and Survey categories. The pronounced variability in SRs reflects the model’s difficulty generating high-quality feedback when learner submissions contain limited detail. This is an issue echoed in the main analysis and consistent with the pedagogical challenges of providing feedback on early-stage work.

Overall, while human raters generally provided higher scores, specific items across all report types and case types demonstrated instances of superior AI performance, indicating that the relative effectiveness of AI versus human feedback is highly dependent on the specific context, dimension, and item being evaluated. This item-level variability also highlights potential focus areas for future prompt refinement or domain-specific tuning.

**Table 4.** Final Reports: Item level difference scores between Human and AI feedback (1-5 scale)

| Case | Quality | Understanding | Persona | Safety | Trust | Case | Quality | Understanding | Persona | Safety | Trust |
| --- | --- | --- | --- | --- | --- | --- | --- | --- | --- | --- | --- |
| Literature Review |  |  |  |  |  | Research Report |  |  |  |  |  |
| ESL1 | 0.67 | 1.00 | 1.00 | -1.20 | 0.00 | ESR1 | 0.75 | 1.00 | 1.33 | 0.00 | 1.50 |
| ESL2 | -0.83 | -0.33 | 0.33 | -0.20 | -1.50 | ESR2 | 0.00 | 0.00 | 0.33 | 0.00 | 0.00 |
| ESL3 | -0.26 | -0.67 | 0.33 | -1.60 | -1.00 | ESR3 | 2.00 | 1.67 | 2.00 | 1.80 | 2.00 |
| ESL4 | 1.83 | 1.33 | 0.67 | 1.80 | 3.00 | ESR4 | 1.00 | 0.67 | 0.33 | 0.20 | 1.50 |
| ESL5 | 0.17 | 0.00 | 0.33 | -1.00 | 0.50 | ESR5 | -0.58 | -0.67 | -0.33 | 0.20 | 0.00 |
| ESL6 | 0.42 | 1.00 | 0.67 | 0.80 | 0.50 | ESR6 | 1.17 | 0.33 | 0.00 | 1.00 | 1.00 |
| ESL7 | 0.17 | 0.00 | 0.67 | -0.60 | 0.50 | ESR7 | -0.75 | -1.00 | -1.00 | -0.20 | -1.00 |
| ESL8 | 0.75 | 0.33 | 0.00 | -0.80 | 1.00 | ESR8 | -1.08 | -1.33 | -0.67 | -0.80 | -0.50 |
| ESL9 | 1.00 | 0.33 | 0.67 | -1.00 | 1.00 | ESR9 | 0.92 | 1.00 | 1.00 | 0.20 | 1.00 |
| ESL10 | 0.08 | -0.67 | 0.00 | -0.20 | -0.50 | ESR10 | 0.00 | 0.00 | 0.33 | 0.00 | 0.50 |
| Quality Improvement |  |  |  |  |  | Survey |  |  |  |  |  |
| ESQ1 | 0.25 | 0.00 | 0.00 | 0.80 | -1.50 | ESS1 | -1.17 | -0.33 | 0.00 | -0.80 | -1.00 |
| ESQ2 | -0.33 | 0.00 | 0.00 | 1.00 | -0.50 | ESS2 | -1.00 | -0.67 | -0.67 | 0.00 | -0.50 |
| ESQ3 | -1.25 | -1.00 | -1.00 | -0.40 | -1.50 | ESS3 | 0.00 | 0.00 | 0.00 | 0.00 | 0.00 |
| ESQ4 | 0.25 | 0.67 | 0.33 | -0.80 | 1.00 | ESS4 | 0.36 | 1.00 | 0.67 | -0.80 | 0.00 |
| ESQ5 | 0.17 | -1.00 | -0.67 | 0.40 | 0.00 | ESS5 | -0.67 | -1.00 | -0.33 | 0.60 | -1.00 |
| ESQ6 | 0.42 | 0.00 | 0.67 | -1.00 | -1.00 | ESS6 | 0.92 | 0.67 | 1.00 | 0.20 | 1.00 |
| ESQ7 | 0.92 | 1.00 | 1.00 | 0.40 | 1.00 | ESS7 | -0.92 | -1.00 | -0.67 | -1.00 | -1.50 |
| ESQ8 | 0.08 | 0.00 | -0.33 | 0.80 | -0.50 | ESS8 | -0.25 | 0.00 | 0.33 | 0.00 | 0.00 |
| ESQ9 | -0.08 | 0.00 | 0.00 | -0.80 | 0.00 | ESS9 | 0.17 | -1.33 | 0.00 | -1.60 | -1.50 |
| ESQ10 | 0.92 | 1.33 | 1.33 | -1.00 | 1.00 | ESS10 | -0.50 | 0.00 | 0.33 | 0.00 | 0.00 |

Note: Negative values indicate higher scores for AI than for Human feedback

**Table 5.** Question and Timeline Reports: Item level difference scores between Human and AI feedback (1-5 scale)

| Case | Quality | Understanding | Persona | Safety | Trust | Case | Quality | Understanding | Persona | Safety | Trust |
| --- | --- | --- | --- | --- | --- | --- | --- | --- | --- | --- | --- |
| Literature Review |  |  |  |  |  | Research Report |  |  |  |  |  |
| EQL1 | 1.50 | 1.00 | 0.33 | 0.00 | 1.50 | EQR1 | -0.25 | 0.00 | 0.33 | 0.75 | 0.00 |
| EQL2 | 0.33 | 0.00 | 1.00 | 2.25 | 0.50 | EQR2 | 3.17 | 4.00 | 3.33 | 0.25 | 3.50 |
| EQL3 | -0.75 | -1.00 | 0.00 | 0.00 | 0.50 | EQR3 | 0.25 | 0.00 | 0.00 | -1.75 | 1.00 |
| EQL4 | 2.00 | 1.33 | 1.33 | 2.00 | 2.00 | EQR4 | -0.42 | -1.00 | -1.00 | 0.00 | -1.50 |
| EQL5 | 1.25 | 2.00 | 1.67 | 1.00 | 2.00 | EQR5 | 1.00 | 1.67 | 2.33 | 0.00 | 1.50 |
| EQL6 | 1.08 | 1.33 | 2.00 | 0.00 | 1.00 | EQR6 | 1.00 | 1.33 | 2.33 | 0.50 | 2.00 |
| EQL7 | -0.67 | -1.33 | -1.00 | 0.00 | -0.50 | EQR7 | 1.25 | 1.33 | 0.67 | 0.00 | 0.50 |
| EQL8 | 1.75 | 1.33 | 1.33 | 1.50 | 2.00 | EQR8 | -1.08 | -0.33 | -0.67 | 0.00 | 0.00 |
| EQL9 | 2.67 | 1.00 | 0.67 | 1.50 | 2.50 | EQR9 | -1.00 | -1.33 | -1.33 | 0.00 | -1.00 |
| EQL10 | 1.75 | 1.67 | 1.00 | 1.25 | 2.00 | EQR10 | 0.67 | 0.33 | 0.00 | 0.00 | 1.50 |
| Quality Improvement |  |  |  |  |  | Survey |  |  |  |  |  |
| EQQ1 | 0.58 | 0.33 | 0.67 | -0.50 | 1.50 | EQS1 | 1.50 | 1.00 | 1.00 | 1.50 | 2.00 |
| EQQ2 | 1.00 | 1.00 | 0.33 | 0.25 | 1.00 | EQS2 | -0.17 | 0.67 | 0.33 | 0.75 | 0.00 |
| EQQ3 | 0.67 | 0.00 | 1.33 | -0.25 | 0.00 | EQS3 | 0.08 | 0.00 | 0.00 | 1.75 | 0.50 |
| EQQ4 | 0.92 | 0.00 | 0.33 | 0.50 | 0.00 | EQS4 | 0.75 | 2.67 | 1.00 | 0.00 | 1.00 |
| EQQ5 | -1.08 | -0.67 | 0.00 | -1.00 | 0.00 | EQS5 | 0.92 | 1.67 | 1.33 | 1.50 | 1.50 |
| EQQ6 | -0.33 | 0.00 | 0.00 | -0.50 | 1.00 | EQS6 | 1.00 | 1.33 | 1.33 | 0.00 | 1.50 |
| EQQ7 | 1.50 | 1.00 | 1.00 | 2.25 | 1.50 | EQS7 | 0.92 | 1.00 | 0.33 | 2.00 | 1.00 |
| EQQ8 | 0.00 | -0.67 | -0.33 | -1.25 | -1.50 | EQS8 | 0.33 | 0.67 | 0.67 | 0.00 | 1.50 |
| EQQ9 | 0.42 | 0.67 | 0.33 | 0.00 | 0.50 | EQS9 | 1.08 | 0.67 | 0.67 | 0.25 | 0.50 |
| EQQ10 | 1.50 | 1.00 | 1.00 | 1.50 | 2.00 | EQS10 | 0.00 | 1.00 | 1.00 | 1.25 | 1.00 |

Note: Negative values indicate higher scores for AI than for Human feedback

**Table 6.** Short Reports: Item level difference scores between Human and AI feedback (1-5 scale)

| Case | Quality | Understanding | Persona | Safety | Trust | Case | Quality | Understanding | Persona | Safety | Trust |
| --- | --- | --- | --- | --- | --- | --- | --- | --- | --- | --- | --- |
| Literature Review |  |  |  |  |  | Research Report |  |  |  |  |  |
| ESL7 | 3.08 | 3.00 | 2.00 | 2.40 | 3.00 | ESR7 | 2.17 | 0.67 | 0.33 | -1.00 | 2.00 |
| ESL2 | 2.25 | 3.67 | 3.00 | 2.60 | 2.50 | ESR6 | 1.67 | 1.33 | 0.67 | 0.40 | 2.00 |
| ESL3 | 1.83 | 2.00 | 1.33 | 1.80 | 2.00 | ESR9 | 1.33 | 1.00 | 1.00 | 1.00 | 1.00 |
| ESL6 | 1.75 | 0.67 | 0.33 | 0.40 | 2.00 | ESR4 | 1.25 | 1.00 | 1.00 | 1.00 | 1.50 |
| ESL8 | 1.00 | 0.67 | 1.00 | 1.00 | 1.00 | ESR5 | 1.17 | 1.00 | 0.67 | 1.60 | 2.00 |
| ESL9 | 0.83 | 0.00 | -0.33 | -0.60 | 0.50 | ESR3 | 0.75 | 0.33 | 0.00 | 0.80 | 1.00 |
| ESL10 | 0.58 | 1.33 | 1.00 | 0.20 | 0.50 | ESR2 | 0.67 | 1.00 | -0.33 | 0.00 | 0.50 |
| ESL4 | 0.50 | 0.33 | 0.67 | 1.00 | 2.00 | ESR1 | 0.25 | 0.00 | 0.33 | -0.40 | 1.00 |
| ESL1 | -0.17 | 0.00 | -1.00 | -0.80 | 0.00 | ESR10 | 0.17 | -0.33 | -1.00 | -2.60 | 0.50 |
| ESL5 | -0.67 | -1.00 | -0.33 | -1.60 | -0.50 | ESR8 | -0.25 | 0.00 | 0.00 | 0.60 | -0.50 |
| Quality Improvement |  |  |  |  |  | Survey |  |  |  |  |  |
| ESQ3 | 2.75 | 2.33 | 2.33 | 2.00 | 3.00 | ESS4 | 2.83 | 3.00 | 3.00 | 1.00 | 2.50 |
| ESQ1 | 2.67 | 2.33 | 2.67 | 1.00 | 2.00 | ESS3 | 2.00 | 1.00 | 0.00 | -0.20 | 2.00 |
| ESQ6 | 2.55 | 3.00 | 2.33 | 2.20 | 1.50 | ESS5 | 1.83 | 1.67 | 1.33 | 2.00 | 2.00 |
| ESQ8 | 2.50 | 3.00 | 2.67 | 2.00 | 3.00 | ESS7 | 1.42 | 1.00 | 1.00 | -0.20 | 1.50 |
| ESQ7 | 1.50 | 1.00 | 1.33 | 0.80 | 1.00 | ESS6 | 1.17 | 0.67 | 0.00 | 1.00 | 2.00 |
| ESQ2 | 1.42 | 1.33 | 0.67 | 1.00 | 1.00 | ESS9 | 0.58 | -0.33 | -0.33 | 0.00 | 1.50 |
| ESQ5 | 1.25 | 1.67 | 1.00 | 0.60 | 3.00 | ESS2 | 0.00 | 0.00 | 0.00 | 0.00 | 0.00 |
| ESQ10 | 1.08 | 3.00 | 2.00 | 1.00 | 2.00 | ESS10 | -0.83 | -1.00 | -1.33 | -0.20 | -1.50 |
| ESQ4 | 0.33 | 1.67 | 1.00 | 1.00 | 0.50 | ESS1 | -1.17 | -1.00 | -1.00 | -1.20 | -1.50 |
| ESQ9 | 0.00 | 0.00 | 0.00 | 0.00 | 0.00 | ESS8 | -1.83 | -2.00 | -2.00 | -2.00 | -2.00 |

Note: Negative values indicate higher scores for AI than for Human feedback

## Appendix II System Prompt

### # Instructions

You are now an expert evaluator for student university projects in the healthcare and medicine fields. Your role is to provide a detailed, constructive evaluation of the student’s project, guiding them through what they did correctly and what could be improved. Your evaluation should focus on clarity, academic rigor, and alignment with the standards found in published healthcare science reports.

- When giving feedback, always refer to specific sections of the report.
- Provide actionable and practical advice to help the student improve their project.
- Use concrete examples to clarify what adjustments the student can make.
- The student will provide you with information related to their healthcare project, classified as a {paper_type}. The evaluation will focus on the following key aspects:

### # PROJECT QUESTION

The question of the student’s project. Assess whether the title and question are clear and aligned with the project’s objectives. The title should reflect the scope of the research accurately and set clear expectations for the reader. Suggest any necessary refinements.

### # METHODOLOGY

A summary of the project’s methodology. Evaluate whether the methodology is appropriate, well-structured, and clearly described. Ensure that the project question is well-supported by the methodology. Offer feedback on areas needing more detail.

Evaluate the following components:

- Objective: Check if the objective is clearly defined and aligns with the research question.
- Design: Review whether the study design is suitable for addressing the research question.
- Setting: Determine if the context in which the project is conducted is appropriate and well-explained.
- Population: Assess whether the target population is clearly defined, including any relevant criteria.
- Intervention: If applicable, verify that the intervention is relevant and appropriately described.
- Data Collection: Evaluate whether the data to be collected is clearly specified and the methods for data collection are suitable for the project.

In addition to these components, it is crucial to evaluate the methodology against the given criteria:

{criteria}{reb}

### # TIMELINE

The timeline for project completion. Ensure that the timeline is realistic given the scope of the project. Check whether the student has accounted for important steps the complexity of the literature search, and time for publication. {timeline} Suggest adjustments if necessary.

### # CLASSIFICATION OF THE PROJECT

The project classification is between a scholarly and a research project. Confirm whether the METHODOLOGY aligns with the project classification selected by the student. Ensure the project fits the criteria for either a scholarly project or a research project. If there is misalignment, suggest how the student could modify their approach or presentation to better fit the selected type.

Scholarly Project:

- Definition: A scholarly project is any written work that is produced by a scholar or researcher for the purpose of contributing to academic knowledge. It is typically published in academic journals, books, or conference proceedings.
- Purpose: The goal of a scholarly project is to provide insights, arguments, or critiques based on careful analysis or review of existing knowledge. It may not always involve new original research but could include theoretical exploration, literature reviews, or essays.
- Examples: Review articles, theoretical papers, opinion pieces, literature reviews.

Research Project:

- Definition: A research project specifically refers to a document that presents the results of original research conducted by the author(s). It involves empirical investigation, data collection, analysis, and interpretation.
- Purpose: The primary objective of a research project is to present new findings or discoveries. It typically includes the research question or hypothesis, methodology, data analysis, and conclusions.
- Examples: Experimental studies, clinical trials, fieldwork reports, quantitative or qualitative studies.

### # PUBLISHING

Whether the student plans to publish the paper. Consider the plan for publication when evaluating the timeline and the project’s methodology. This information will also help you for the timeline information to see if it is realistic.

|  | {criteria} | {reb} |
| --- | --- | --- |
| Survey | - Describe the source of the questions (e.g., validated survey from literature)<br>- Describe the type of questions (e.g., multiple-choice, open-ended, etc.)<br>- Describe how you will administer the survey.<br>- Describe your sample and sample size.<br>- Describe your recruitment strategy.<br>- Describe the data you will collect.<br>- Describe your data analysis plan and where the data will be stored. | The student should apply for Research Ethics Board (REB) when:<br>- Generating new knowledge where intent is to generalize findings<br>- Information that the student is gathering is not anonymous or publicly available<br>- It is a publication & presentation<br>- Implementing an intervention (e.g. procedure, medication or service)<br>- It involves human subjects or their health information<br>Types of REB:<br>- The primary REB (aka Board of Record) should be selected based on the student's affiliation (e.g., research conducted by TOH/OHRI personnel requires OHSN-REB) |
|  |  | approval, regardless of where the research is taking place).<br>- Other REB approvals' requirement is based on where the research and recruitment of participants is taking place.<br>REB Approval Timeline:<br>- Projects that undergo a delegated/expedited review are faster (about 6 to 8 weeks)<br>- Low risk to the population involved<br>- Generally involve data extraction, surveys, focus groups, etc.<br>- Projects with Ottawa Health Science Network (OHSN) with an IRIS Ethical Application (CRRF) (about 6 to 8 weeks)<br>- Projects with Bruyère with a COREB Common application form (about 6 to 8 weeks)<br>- Projects with Montfort with a COREB Common application form (about 2 to 4 weeks)<br>- Projects with uOttawa with eReviews (about 4 to 6 weeks) |
| Literature Review | - Provide details on your search, selection, and synthesis<br>– how will you find papers, select which to keep, and summarize the results?<br>- Include a list of key search terms and MeSH terms.<br>- Define your inclusion/exclusion criteria.<br>- List potential databases for your search. | n/a |
| Research Study | - Describe the study design (e.g., pre-post study design, prospective study, etc.)<br>- Describe your sample and sample size.<br>- Describe your inclusion/exclusion criteria. | The student should apply for Research Ethics Board (REB) when:<br>- Generating new knowledge where intent is to generalize findings |
|  | - Describe your recruitment strategy (if applicable)<br>- Define your outcome measures (ensure they are linked to your project question)<br>- Describe the data you will collect.<br>- Describe the data analysis plan and where the data will be stored. | - Information that the student is gathering is not anonymous or publicly available<br>- It is a publication & presentation<br>- Implementing an intervention (e.g. procedure, medication or service)<br>- It involves human subjects or their health information<br>Types of REB:<br>- The primary REB (aka Board of Record) should be selected based on the student's affiliation (e.g., research conducted by TOH/OHRI personnel requires OHSN-REB approval, regardless of where the research is taking place).<br>- Other REB approvals' requirement is based on where the research and recruitment of participants is taking place.<br>REB Approval Timeline:<br>- Projects that undergo a delegated/expedited review are faster (about 6 to 8 weeks)<br>- Low risk to the population involved<br>- Generally involve data extraction, surveys, focus groups, etc.<br>- Projects with Ottawa Health Science Network (OHSN) with an IRIS Ethical Application (CRRF) (about 6 to 8 weeks)<br>- Projects with Bruyère with a COREB Common application form (about 6 to 8 weeks)<br>- Projects with Montfort with a COREB Common application form (about 2 to 4 weeks) |
|  |  | - Projects with uOttawa with eReviews (about 4 to 6 weeks) |

## Appendix III Code

GitHub link: [to add prior to submission]

## Notes

### Competing Interest Statement

The authors have declared no competing interest.

### Author Declarations

This project falls under TCPS2 Article 2.5 and, as such, did not require Research Ethics Board (REB) review, as confirmed by the University of Ottawa REB during an initial review for exemption and reported on August 29, 2024.

### Summary of Updates

Typo in author credentials corrected

